# A Bayesian System to Track Outbreaks of Influenza-Like Illnesses Including Novel Diseases

**DOI:** 10.1101/2023.05.10.23289799

**Authors:** John M. Aronis, Ye Ye, Jessi Espino, Harry Hochheiser, Marian G. Michaels, Gregory F. Cooper

**Affiliations:** Department of Biomedical Informatics, University of Pittsburgh, 5607 Baum Blvd, Suite 500, Pittsburgh, PA 15206-3701; Department of Pediatrics, School of Medicine, University of Pittsburgh, S530 Alan Magee Scaife Hall, 3550 Terrace Street, Pittsburgh, PA 15261

## Abstract

It would be highly desirable to have a tool that detects the outbreak of a new influenza-like illness, such as COVID-19, accurately and early. This paper describes the *ILI Tracker* algorithm that first models the daily occurrence of a set of known influenza-like illnesses in a hospital emergency department using findings extracted from patient-care reports using natural language processing. We include results based on modeling the diseases influenza, respiratory syncytial virus, human metapneumovirus, and parainfluenza for five emergency departments in Allegheny County Pennsylvania from June 1, 2010 through May 31, 2015. We then show how the algorithm can be extended to detect the presence of an unmodeled disease which may represent a novel disease outbreak. We also include results for detecting an outbreak of an unmodeled disease during the mentioned time period, which in retrospect was very likely an outbreak of Enterovirus D68.

## 1 Introduction

A set of respiratory viruses is responsible for annual and oftentimes overlapping outbreaks in human populations. This overlapping disease activity confounds diagnosis and treatment of patients presenting with influenza-like illnesses and the associated high caseloads stress the clinical and logistical capacity of the healthcare system. Thus, accurately detecting and tracking overlapping outbreaks due to these viruses is an important task with public health implications and clinical repercussions for those at high risk [1, 2, 3, 4, 5]. The ideal surveillance system will notice an outbreak after just a few cases that may be distributed acsross several shifts at multiple hospitals. An individual physician may see just one or two cases, which might seem inconsequential to them and not worthy of mention, but an automated surveillance system, such as we describe here, can gain statistical power and timeliness by aggregating data across an entire region. The proposed system can harness the entire set of patients and their symptoms who present to all the emergency departments in a region. By harnessing the sheer volume of this information, it may recognize cases and patterns that would elude human observers early in the outbreak, and thus, serve as a sentinel to detect and characterize outbreaks early.

In addition to detecting and tracking known viruses of concern, the world is also faced with the emergence of novel viruses (or the re-emergence of previously quiescent viruses) and the diseases that they cause, as evidenced by the appearance of the COVID-19 worldwide pandemic. Early detection and tracking of a novel or re-emerging outbreak disease can be critical in informing both the care of individual patients and the decisions made by public health officials. While we hope to someday prevent the emergence of pathological viruses before they strike the human population [6], a more realistic goal for the near-term is early detection and tracking [7]. Modeling known diseases that we expect to see provides a useful background against which to detect the emergence of new diseases that have a different clinical or epidemiological presentation.

This paper describes the ILI Tracker algorithm which tracks the daily occurrence of a set of modeled influenza-like illnesses (ILIs) in a hospital emergency department (ED) using natural language processing (NLP) on patient care reports. A set of clinical findings are extracted from full-text patient care reports that are available at the time of care or shortly thereafter. These findings are used by machine learning algorithms to learn probabilistic models of a set of diseases. These models are needed to determine the likelihood of each disease for each patient [8]. These likelihoods are then used to compute the expected rate of each disease each day. ILI Tracker also analyzes whether recent patient cases in the ED are not well explained by its models of known diseases. If so, it suggests the possible presence of a novel outbreak disease in the population.

The remainder of this paper describes the ILI Tracker algorithm in detail, preliminary experiments with it for the modeled diseases influenza, respiratory syncytial virus (RSV), human metapneumovirus (hMPV), and parainfluenza (PIV) for five EDs in Allegheny County Pennsylvania from June 1, 2010 through May 31, 2015. We also present a preliminary investigation of an algorithm based on ILI Tracker for detecting the presence of an unmodeled disease.

## 2. The Algorithm

To diagnose a patient, a clinician must account for the findings of that patient as well as the prevalence of various diseases in the community. For instance, given a patient with a fever and cough, a high rate of influenza in the population will elevate the probability the patient has influenza, whereas a high rate of COVID-19 will elevate the probabilty of COVID-19. The situation becomes more complicated when multiple viruses (with similar or overlapping symptoms) are circulating in the environment for which the rate of each must be accounted. Bayesian inference provides a principled way to do this.

If *Pr*(*dx*_*i*_|*findings*) is the probability that a patient has disease *dx*_*i*_ given their *findings*, and *Pr*(*dx*_*i*_) is the rate of that disease in the population at that time, then:

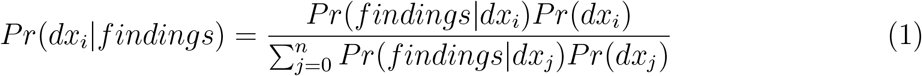

The denominator is a sum over all the diseases we are modeling and is a normalizing factor, so we have:

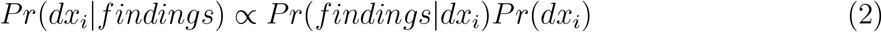

That is, the probability the patient has disease *dx*_*i*_ given their *findings* is proportional to the product of the likelihood of their *findings* given *dx*_*i*_ times the prior probability of the *dx*_*i*_.

Using this formulation, we can compute the probability of each disease for each patient. The expected number of patients with each disease is the sum over the probability each patient has that disease. For example, suppose there are 50 patients and 20 have a 0.1 probability of influenza, while 30 have a 0.2 probability. Then the expected number of patients with influenza is 20 × 0.1 + 30 × 0.2 = 8. Given the expected number with each disease, we can compute the expected proportion of each disease. In the example above the expected proportion of patients with influenza would be 8*/*50 = 0.16.

The ILI Tracker algorithm combines the above steps to compute the expected proportion of each disease each day: start with prior probabilities for each disease on the first day, compute the expected number of patients with each disease as above, then use the proportion of each disease as the prior probability of each disease on the next day. Continue this process day-by-day. The remainder of this section provides the technical details of this algorithm.

We first introduce some notation. Let *days* be the sequence of days under consideration, *pts*(*d*) the number of patients on day *d, D*(*p, d*) the set of findings (‘data’) for patient *p* on day *d*, and 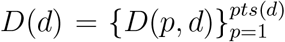 is all of the data for the patients on day *d*. We assume there is a set of modeled diseases *Dx* = {*dx*_0_, *dx*_1_, …, *dx*_*n*_} where {*dx*_1_, …, *dx*_*n*_} are the diseases of interest and *dx*_0_ denotes *other* known diseases. Here, *other* represents a large set of known diseases including trauma, cardiac events, diabetic emergencies, etc., that can occur and are not one of the other *n* modeled diseases.

For a patient *p* on day *d* with findings *D*(*p, d*) we can calculate the probability they have a particular disease *dx*_*i*_:

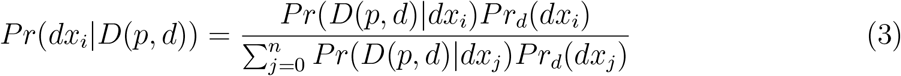

where *Pr*_*d*_(*dx*_*j*_) is the prior probability of *dx*_*j*_ on day *d* and *Pr*(*D*(*p, d*)|*dx*_*i*_) is the likelihood of patient *p*’s findings given they have disease *dx*_*i*_. We describe how we compute each of these quantities below. We compute the expected number of patients with each *dx*_*i*_ on day *d* as:

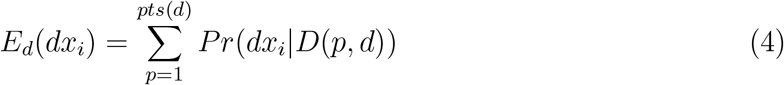

We can now estimate the posterior probability of each disease on day *d*:

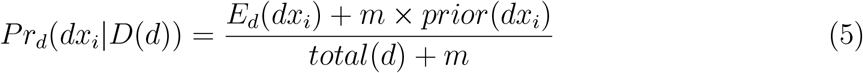

where *m* is the so-called *equivalent sample size, total*(*d*) is the total number of patients on day *d*, and *prior*(*dx*_*i*_) is the prior probability of disease *dx*_*i*_. The terms *m* and *prior*(*dx*_*i*_) in Equation 5 provide smoothing of the estimate that avoids relying too heavily on small values of *E*_*d*_(*dx*_*i*_) and *total*(*d*) by augmenting the data with an additional *m* virtual patients with diseases distributed according to *prior*(*dx*_*i*_). We specify *m* and *prior*(*dx*_*i*_) below. We then make the disease priors for day *d* + 1 equal to the disease posteriors for day *d*.

We compute the expected number of patients with each disease day-by-day. Set each *Pr*_0_(*dx*_*i*_) to initial values. Then for each day *d* ≥ 1:

1. Compute *Pr*(*dx*_*i*_|*D*(*p, d*)) for each *p* ∈ *Pts*(*d*) using Equation 3.
2. Compute *E*_*d*_(*dx*_*i*_) for each *dx*_*i*_ using Equation 4.
3. Compute *Pr*_*d*_(*dx*_*i*_|*D*(*d*)) for each *dx*_*i*_ using Equation 5
4. Set *Pr*_*d*+1_(*dx*_*i*_) = *Pr*_*d*_(*dx*_*i*_|*D*(*d*)) for each *dx*_*i*_. Steps 1-4 are repeated for each successive day.

The prior probabilities on June 1, 2014 for influenza, RSV, hMPV, and PIV were set to 0.033, 0.035, 0.005, and 0.003, respectively. We determined these numbers by running ILI Tracker on the data for March 1 through May 31 of 2014 with priors on March 1 of 0.05, 0.05, 0.05, and 0.05, and using the resulting posterior probabilities from May 31 as the prior probabilities for June 1. The equivalent sample size was set to 10, and the *Pr*_0_(*dx*_*i*_) values were set to the prior probability of each modeled disease on June 1 as described above.

## 3 Detecting the Presence of Unmodeled Diseases

As mentioned above, by modeling the diseases we expect to see in the ED we can better detect novel diseases. The remainder of this section specifies how this is performed.

We can regard the output of ILI Tracker as a model of the types of patients who are in the ED each day. ILI Tracker assumes the presence of a fixed set of diseases that can be modeled using Bayesian networks with specified findings (see Table 1). If this assumption is satisfied then the model should explain the evidence (the patients and their findings) well, and the probability of the data given the diseases in an outbreak will be relatively high.

**Table 1:** Clinical features and their ordering used by the K2 algorithm.

|  |  |
| --- | --- |
| 1 lab positive vrp - pathogen not specified | 41 cervical lymphadenopathy |
| 2 parainfluenza | 42 infiltrate |
| 3 metapneumovirus | 43 productive cough |
| 4 abdominal pain | 44 staccato cough |
| 5 anorexia | 45 grunting |
| 6 apnea | 46 sore throat |
| 7 arthralgia | 47 nasal flaring |
| 8 bronchiolitis | 48 other cough |
| 9 bronchitis | 49 decreased activity |
| 10 chest pain | 50 lab order (nasal swab) |
| 11 conjunctivitis | 51 highest measured temperature |
| 12 croup | 52 crackles |
| 13 cyanosis | 53 respiratory distress |
| 14 diarrhea | 54 influenza-like illness |
| 15 dyspnea | 55 poor feeding |
| 16 weakness or fatigue | 56 toxic appearance |
| 17 headache | 57 hypoxemia (SpO2 on room air < 90%) |
| 18 hemoptysis | 58 other abnormal breath sounds |
| 19 hoarseness | 59 nonproductive cough |
| 20 rigor | 60 runny nose |
| 21 stuffy nose | 61 acute onset |
| 22 nausea | 62 lab testing ordered (rsv) |
| 23 pharyngitis diagnosis | 63 respiratory pathogens panel ordered) |
| 24 other pneumonia | 64 ill-appearing |
| 25 viral pneumonia | 65 reported fever |
| 26 rales | 66 chest wall retractions |
| 27 rhonchi | 67 bilateral acute conjunctivitis |
| 28 seizure | 68 lab positive rsv |
| 29 stridor | 69 lab positive strep a |
| 30 upper respiratory infection | 70 lab positive rhinovirus |
| 31 viral syndrome | 71 lab positive parainfluenza |
| 32 vomiting | 72 lab positive adenovirus |
| 33 wheezing | 73 lab positive enterovirus |
| 34 chills | 74 lab testing ordered (influenza) |
| 35 malaise | 75 lab positive influenza |
| 36 myalgia | 76 lab positive hmpv |
| 37 tachypnea | 77 poor response of fever to antipyretics |
| 38 paroxysmal cough | 78 pharyngitis on exam |
| 39 abdominal tenderness | 79 abnormal chest radiograph findings |
| 40 barking cough |  |

If any of these assumptions are violated—in particular, if there are patients with a novel, unmodeled disease—the probability of the data given the model produced by ILI Tracker will likely be reduced compared to a previous period of time when only modeled diseases were present in the ED. If we track the probability of the data given the output of ILI Tracker, a decrease in this daily probability may signal the presence of an unmodeled disease. An unmodeled disease may be a novel disease or a re-emergent disease that we are not currently modeling.

The day-to-day probabilities for each disease computed by ILI Tracker can be used to perform a posterior predictive check by computing the likelihood of the data each day given the output of ILI Tracker. Specifically, we can compute the likelihood of the data given the output of ILI Tracker each day:

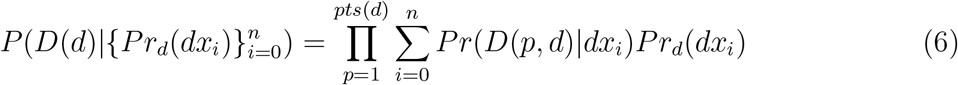

where 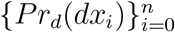 is the set of the prior probabilities of each disease on day *d* computed by ILI Tracker.

Let the null hypothesis be that the likelihood given by Equation 6 for the current day is the same as or greater than the likelihoods for all previously monitored days, up to 100 days. We compute as follows a daily empirical *p*-value, *p*_*d*_, for current day *d* as follows:

1. Compute 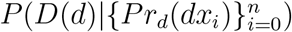
2. Compute 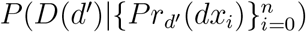 for *d*^′^ = *max*(0, *d* − 100), …, *d* − 1
3. Set *p*_*d*_ to the fraction of times 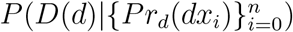 is greater than the values computed in step 2.

That is, we compute the likelihood of the data for the previous (up to) 100 days, then compare the likelihood of the data on day *d* to those values. We say that day *d* is *unusual* if *p*_*d*_ ≤ 0.01. A sequence of unusual days, each with *p*_*d*_ ≤ 0.01, may signify an outbreak.

## 4 Data and Modeling

The training dataset consisted of emergency department (ED) encounters at five University of Pittsburgh Medical Center (UPMC) hospitals from June 1, 2010 through May 31, 2014, including 815 influenza, 414 respiratory syncytial virus (RSV), 198 human metapneumovirus (hMPV), 100 parainfluenza (PIV), and 59,428 “other” visits. We labeled as influenza those patient encounters with a positive laboratory test for influenza by polymerase chain reaction (PCR), direct fluorescent antibody (DFA), or viral culture. We use similar criteria for labeling patient cases with RSV, hMPV, and PIV. For training purposes, we excluded cases that have positive laboratory results for more than one virus. The 59,428 *other* visits were defined as visits in August 2010, 2011, 2012, or 2013, which did not have any influenza, RSV, hMPV, PIV laboratory tests performed. We use August, because relatively few outbreaks of these four diseases occur during that month. The testing dataset consisted of ED encounters from June 1, 2014 through May 31, 2015.

We used the Topaz parser [9] to extract clinical findings and their values. Clinical notes were used as the input for the Topaz parser, which generated a collection of clinical findings and their corresponding values. Among these findings, the highest measured temperature was classified into the four categories high grade (≥ 104.0^◦^F/40.0^◦^C), low grade (100.4^◦^F-103.9^◦^F/38.0^◦^C-39.9^◦^C), inconsequential (*<* 100.4^◦^F/38.0^◦^C), and unknown. Each of the other findings took the values present, absent, or unknown. The designation of “absent” indicated that the clinician had reported the finding as being absent (e.g., “patient denies sore throat”). We discarded findings whose information gain scores (regarding disease diagnosis) is zero. Because there is testing bias for ILIs across age groups, the age feature was not included. Table 1 lists clinical findings coded by Topaz.

We developed five disease models (i.e., influenza, RSV, hMPV, PIV, and other) using the same search process to find each Bayes Network structure as follows. We first initiated a Naive Bayes network structure where all clinical features have an arc from the disease node. We then used the K2 learning algorithm [10] to identify additional arcs among the clinical feature nodes which were assigned an arbitrary order. The search was based on the K2 Bayesian score with a restriction that each clinical feature could have at most two parents beyond the disease node. After finding a network structure, we calculated conditional probabilities in the network model. For instance, Figure 1 shows relationships among the top 20 features (according to information gain) of the RSV model [11].

**Figure 1:**
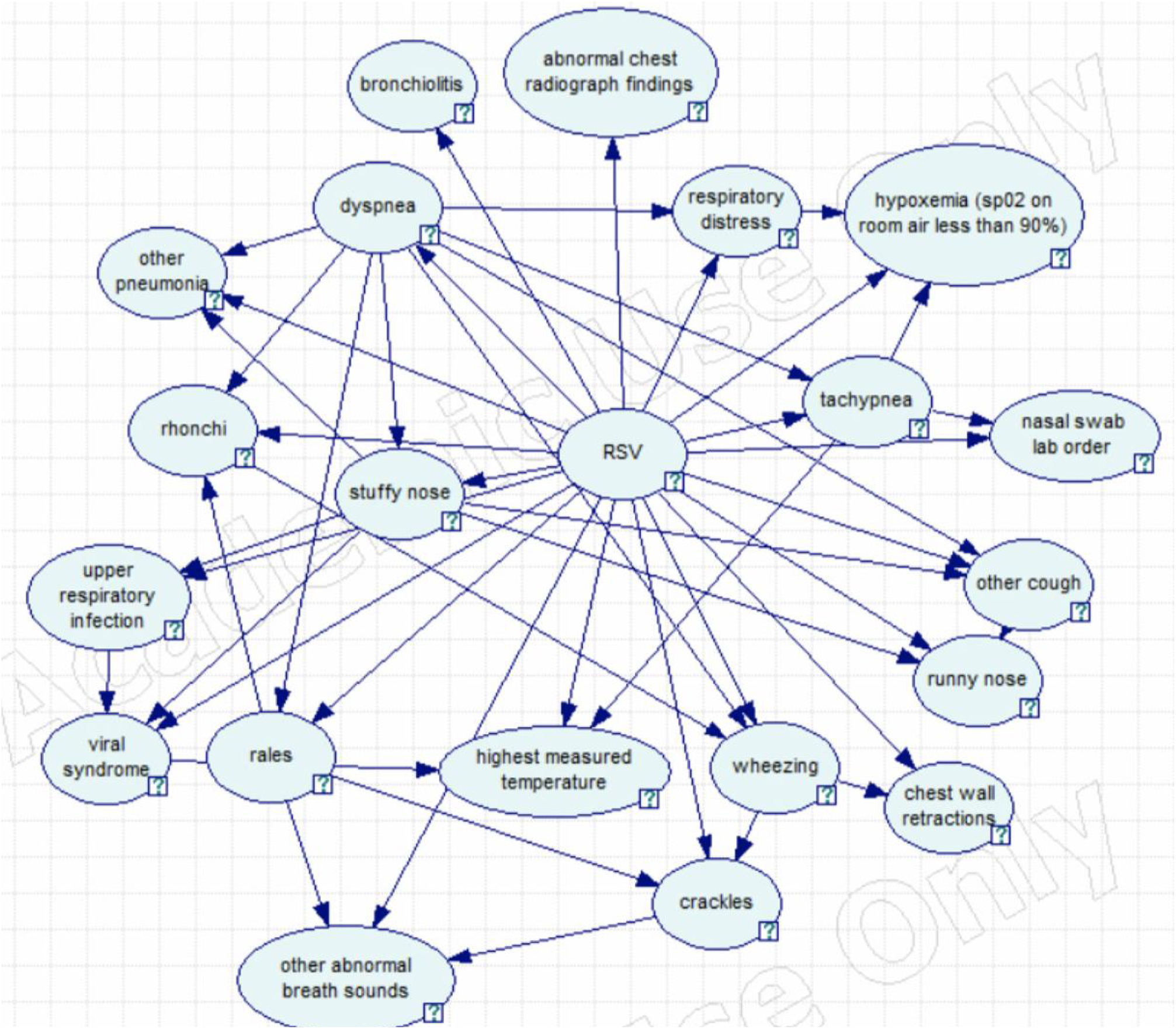
The twenty most informative findings in the RSV model.

Table 2 shows the performance of the five models we developed when tested on one year of data (June 1, 2014 through May 31, 2015). We used the area under the receiver operating characteristic curve (AUC) [12] as the measure of discrimination performance. In testing the performance of the influenza model, the disease-positive group consists of patient cases that have positive laboratory results for influenza. The negative test group consists of cases that either have negative laboratory results for influenza or have not had any laboratory tests performed for influenza. It is likely, however, that some of the patient cases without any laboratory tests for influenza will have influenza, which we would expect to have reduced the AUC reported below. An analogous situation exists for testing the RSV, hMPV, and PIV models.

**Table 2:** Performance of the five developed näive Bayes network disease models.

| Disease | Positive and Negative for training | Positive and Negative for testing | AUC |
| --- | --- | --- | --- |
| Influenza | 815 vs. 60,140 | 654 vs. 266,357 | 0.88 |
| RSV | 414 vs. 60,541 | 449 vs. 266,562 | 0.92 |
| hMPV | 198 vs. 60,757 | 149 vs. 266,862 | 0.91 |
| PIV | 100 vs. 60,855 | 205 vs. 266,806 | 0.89 |
| other | 59,428 vs. 1,527 | 265,588 vs. 1,423 | 0.90 |

When testing the model of *other* disease, the negative test group includes cases that have at least one positive laboratory test result for influenza, RSV, hMPV, or PIV. The positive test group consists of cases that either have negative laboratory results for all four respiratory diseases or have not had any of those tests performed. It is likely, however, that some of the cases without any tests performed will have one or more of the four respiratory diseases. Given the considerations in this and the previous paragraph, the reported AUCs in Table 2 are likely to represent lower bounds on performance that would be obtained if the test case labels were more accurate.

## 5 Experimental Results

This section reports the results we obtained in applying ILI Tracker to the data described above to estimate over time the presence of the outbreak diseases we modeled. We also used our analysis of those known outbreak disease models to predict the presence of a novel disease in the population, which in retrospect was likely due to Enterovirus D68, based on CDC reports for that period.

### 5.1 Tracking of Known Diseases

Figure 2 shows the results of running ILI Tracker for the period June 1, 2014 through May 31, 2015. The red lines are the daily number of laboratory-confirmed cases and the blue lines are the expected number of patients computed by ILI Tracker each day. The ILI-expected-case predictions appear to be correlated with the number of positive laboratory results for those diseases. We computed the correlation between the daily number of expected and confirmed cases for each disease. Table 3 shows the Pearson and Spearman *r* and *p* values, respectively. Across the four modeled diseases, the peak days predicted by ILI Tracker were close to the peak days according to laboratory confirmed cases. The peak days of the seven day moving averages differed by six days for influenza, six days for RSV, zero days for hMPV, and four days for PIV.

**Table 3:** Comparison of ILI Tracker and confirmed cases from June 1, 2014 through May 31, 2015.

| Disease | Pearson $r$ ($p$) | Spearman $r$ ($p$) |
| --- | --- | --- |
| influenza | 0.81 ($< 0.001$) | 0.63, ($< 0.001$) |
| RSV | 0.66 ($< 0.001$) | 0.64, ($< 0.001$) |
| hMPV | 0.72 ($< 0.001$) | 0.65, ($< 0.001$) |
| PIV | 0.51 ($< 0.001$) | 0.52, ($< 0.001$) |

**Figure 2:**
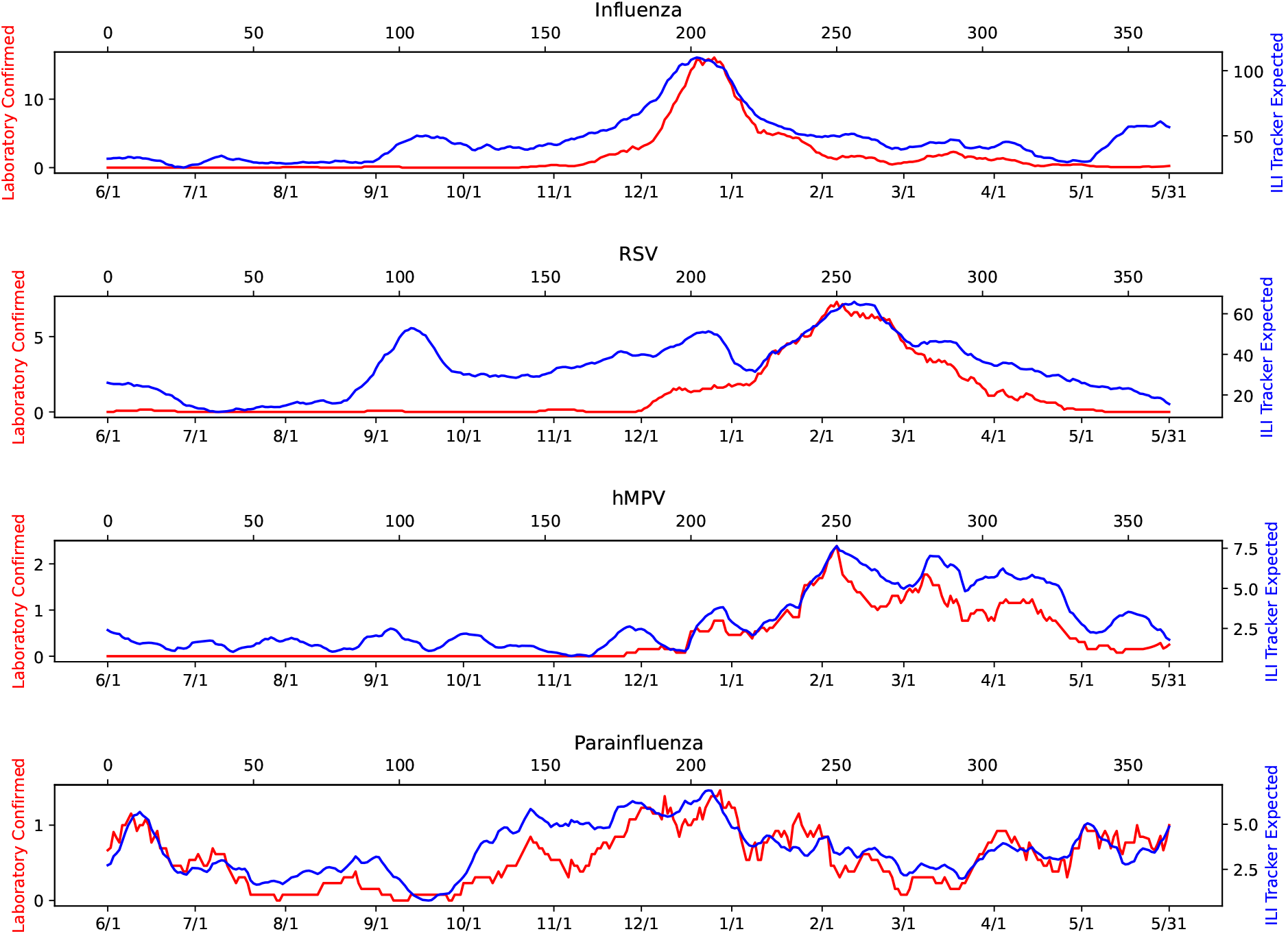
Expected and confirmed cases for June 1, 2014 through May 31, 2015 (seven day moving average).

### 5.2 Putative Detection of a Novel Disease

Figure 3 shows the daily empirical p values for June 1, 2014 through May 31, 2015. The horizontal red line indicates *p* = 0.01 which is our threshold for unusual. On August 25 of 2014, ILI Tracker signaled a highly unusual day. Although a single unusual day is not necessarily the beginning of an outbreak, it may warrant further investigation. Eight of the ten most unusual patients on August 25 (that is, patients with very low probability findings given the expected prevalence of modeled diseases in the ED) showed signs of a respiratory illness. Starting on September 2, ILI Tracker noted a streak of unusual days with the most unusual patients again showing signs of a respiratory illness. By September 9, there were sufficient data to characterize these anomalies in terms of the most prevalent findings. Given the expected number of patients with each modeled disease computed by ILI Tracker on a given day, and the rate of specific findings for the individual viruses, we computed the expected number of each finding per day and compared them to the actual number present. If patients with a novel disease are in fact present in the ED, we expect the actual number of findings in the data characteristic of the novel disease to exceed the expected number of those findings (based on the assumption that only modeled diseases are present). Table 4 shows the top ten most excessive findings for one week starting September 2. Figure 4 tracks the daily occurrence of these findings during July through October 2014. There was a clear increase in their frequency starting in the latter part of August.

**Table 4:** Top 10 excess findings from September 2 through 8, 2014.

|  |
| --- |
| wheezing |
| upper respiratory infection |
| other cough |
| viral syndrome |
| runny nose |
| stuffy nose |
| tachypnea |
| sore throat |
| respiratory distress |
| chest wall retractions |

**Table 5:** Top 10 excess findings from December 1 through 7, 2014.

|  |
| --- |
| other cough |
| viral syndrome |
| stuffy nose |
| upper respiratory infection |
| runny nose |
| sore throat |
| wheezing |
| influenza-like illness |
| tachypnea |
| other pneumonia |

**Figure 3:**
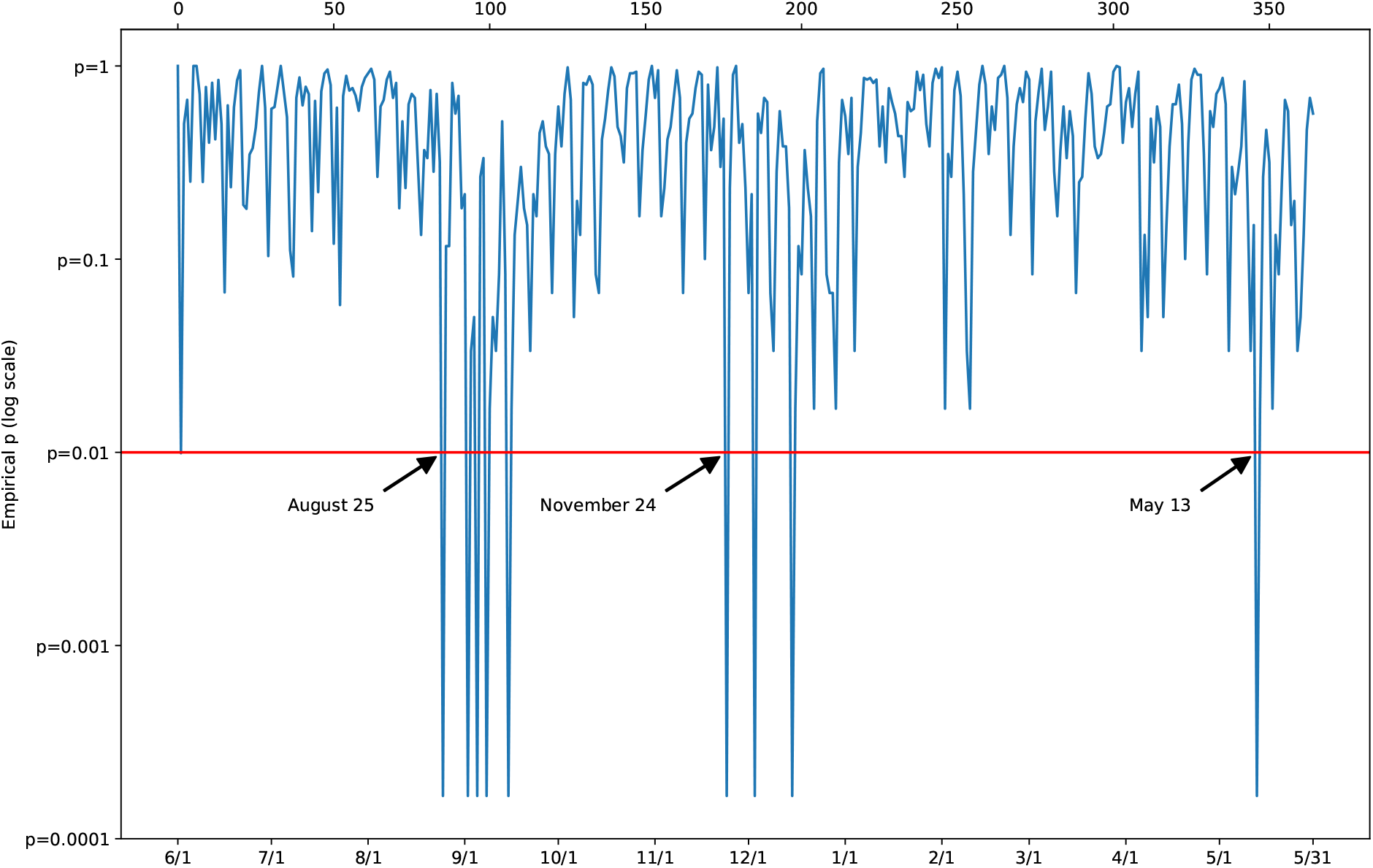
Empirical *p* values for June 1, 2014 through May 31, 2015.

**Figure 4:**
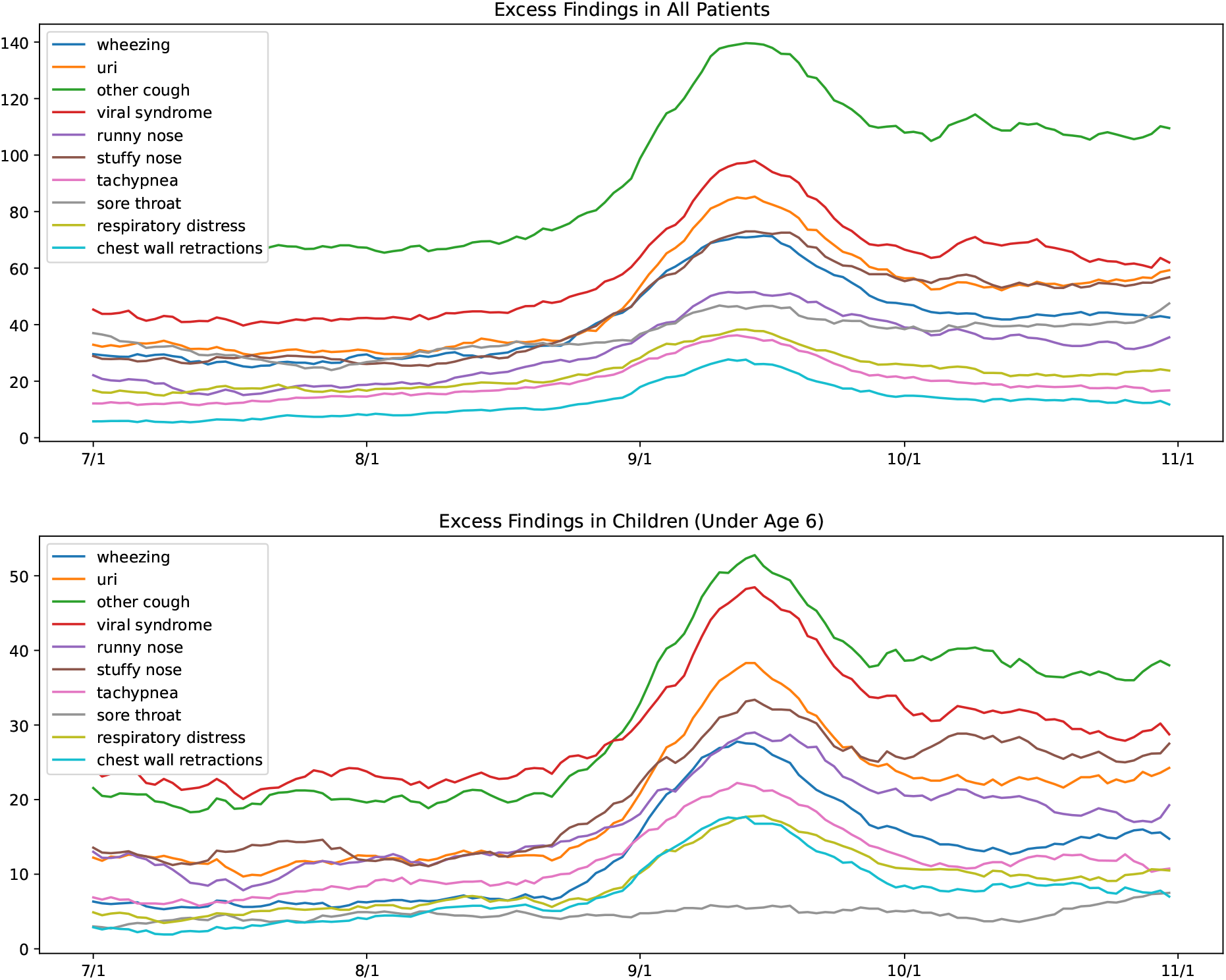
Daily absolute counts of the top ten excess findings July through October 2014.

On November 24 of 2014, ILI Tracker again signaled a day with highly unusual patient findings. This was a weaker signal (see Figure 3) than the signal in late August and early September. Nine of the ten most unusual patients showed signs of a respiratory illness. Figure 5 shows the top ten most excessive findings for the week starting December 1. ILI Tracker also noted an isolated unusual day on May 13, 2015.

### 5.3 Runtime Analysis

We construct disease models once each year (unless there is reason to believe the virus has changed). It took less than one minute using a computer with two processors each with six 1.6GHz cores to construct all the disease models.

Daily processing occurs in three phases: feature extraction from patient care reports, computation of disease likelihoods for each patient, and computation of the expected number of each modeled disease and *p*-value of the data for that day.

There were about 700 patients each day. Feature extraction typically took about five minutes each day using a computer with four 3.6GHz processors. Computation of disease likelihoods typically took less than one minute for the entire set of patients on a given day using the mentioned computer. Given the likelihoods for each patient, the ILI Tracker algorithm takes less than ten seconds to compute the expected number of each modeled ILI and *p*-value of the data each day using a computer with four 2.5GHz cores. Thus, total time to run ILI Tracker per day on a desktop computer is less than ten minutes.

We note that feature extraction and computation of disease likelihoods for each patient care report is independent of the others. Thus, additional healthcare facilities can be added to our surveillance system and each can process their patient care reports locally, with little increase in overall runtime. The runtime of the ILI Tracker algorithm is *O*(*P* × *D* × *F*) each day, where *P* is the number of patients, *D* is the number of diseases, and *F* is the number of features. Processing time increases linearly as patients, diseases, or features are added, and thus, the algorithm is scalable.

## 6. Discussion

The performance of ILI Tracker during a one-year period was moderate for tracking PIV, strong for tracking RSV and hMPV, and very strong for tracking influenza, as measured using Pearson correlation [13] between the tracking and the laboratory confirmed cases. Using Spearman correlation [14], ILI Tracker’s performance was moderate for tracking PIV and strong for tracking hMPV, RSV, and influenza. Interestingly, RSV shows a sizeable peak in September that is not accompanied by an increase in laboratory confirmed cases. We next discuss why we believe this occurred.

During the summer and fall of 2014, the Center for Disease Control and Prevention (CDC) identified an outbreak of Enterovirus D68 (EV-D68) especially among children during this period [15]. Symptoms of EV-D68 include wheezing, difficulty breathing, runny nose, sneezing, cough, body aches, and muscle aches. Acute flaccid myelitis, which is a rare but particularly severe neurologic consequence of EV-D68 leading to paralysis [16], was not included in the symptoms that Topaz represents.

### If ILI Tracker had been in operation during 2014 it would have signaled a statistical anomaly among patients in late August and provided a set of patients for further investigation

Based on clinical judgement, these patients could be assessed, tested, and possibly isolated. In some cases, samples may be obtained for rapid sequencing. By early September, ILI Tracker could also have provided a preliminary clinical description and timeline of a cohort of unusual patients (Table 4 and Figure 4), who turn out to have findings consistent with an outbreak of EV-D68. In the future, such information could help clinicians and public health officials to detect, isolate, characterize, and identify such a novel disease early in the outbreak.

A putative outbreak of an unmodeled disease also appears to have occurred in November 2014 during outbreaks of both RSV and influenza. Although these patients could easily be lost among the large number of patients with RSV and influenza, they are statistically unlikely to be those known diseases, according to the analysis by ILI Tracker. Although this was a weak signal, it was statistically significant enough to warrant further investigation. Again, if ILI Tracker had been in operation at that time it would have identified patients for further evaluation. The source of that putative outbreak remains an open question.

The unusual day detected on May 13, 2015 is likely an isolated incident. Statistically, we should expect such incidents to occur occasionally. ILI Tracker would provide a set of candidate patients for consideration by clinicians and public health officials in the region.

## 7 Related Work

The problem of detecting, characterizing, and forecasting single outbreaks of the common viruses (such as influenza) has been well-studied for over two decades. Some of the best algorithmic methods use mechanistic transmission models (e.g., SIR or SEIR compartment models [17]) and data in the form of univariate time series, weekly counts of diagnostic test results, weekly cases of influenza-like illnesses (ILI), and counts of influenza-related internet queries, for example. Forecasts are made by projecting the best model, or a weighted sum of models, into the future [18, 19, 20, 21, 22]. The research reported in [23] is unique in that it uses humans to extend a trajectory into the future, and therefore—in principle—can indirectly use any information the users are aware of. These methods depend on counts of deterministic events (such as lab tests or ILI counts according to the CDC) and are limited in expressiveness of the information they use. As such, they have limited ability to quickly adapt to novel diseases, whose definitions will not be available early in the outbreak. In our previous work [8, 24, 25, 26] we developed an integrated approach to disease surveillance, including multiple diseases, that starts with the full text of patient-care reports and uses NLP to extract findings (including significant negatives) of each patient seen in a healthcare facility. *Full-text reports are of particular importance because they are often the only record of symptoms and signs, which form the basis for the early detection and characterization of disease outbreaks*.

Research on detecting outbreaks, especially of unmodeled diseases, has relied on a variety of methods and data sources. The simplest *univariate* outbreak detection algorithms (e.g. [27]) track a time-series of a single value, such as emergency department visits or thermometer sales, and look for significant deviations from a baseline level of expected activity. *Multivariate* systems (e.g. [28]) combine several indicators into a single compound indicator in seeking to increase performance. The WSARE (*What’s Strange About Recent Events*) system [29] represents the joint distribution of patient data with a Bayesian network that includes several *environmental attributes* and patient attributes including *reported symptom* (which is similar to a patient’s chief complaint). The network is conditioned on the current values of the environmental attributes to create a conditional joint distribution of response variables for the current day. Thus, the conditional joint distribution represents what would be expected for the current day if there are no outbreaks of new diseases.

A patient’s chief complaint is a concise statement of the symptom or problem that caused the patient to seek medical care. Because patients’ chief complaints are recorded upon admission to a healthcare facility they can provide timely information about disease outbreaks. [30] monitors chief complaints from the current time block to find terms that are anomalous relative to a large baseline (according to Fisher’s exact test applied to counts of each word). The MUSES system [31, 32, 33] uses a semantic scan to infer a set of topics from free-text chief complaints. The method does not rely on existing syndromes or pre-defined illness categories and can recognize one-time events or novel diseases. By comparing topics from recent data to past data the system can recognize anomalous events. By including feedback from a practitioner the MUSES system can de-emphasize uninteresting events.

The DUDE [34] system builds a probabilistic model of normal (baseline) ILI activity using a large set of patient findings extracted from patient-care reports with natural language processing. It then looks for statistically significant deviations from baseline normal activity. Thus, the DUDE system does not rely on just a small set of findings as might be extracted from patients’ chief complaints. By removing cases of known kinds of ILI (such as influenza, RSV, etc.) from the input data, DUDE can recognize new, emergent kinds of ILI.

The DUDE system does not model known kinds of ILI—its input data is just those patients who test negative for all known ILIs. However, throwing away the majority of patients limits its ability to recognize anomalies quickly. ILI Tracker uses the findings for the entire set of patients who present to emergency departments in a region. By modeling known diseases and accounting for their presence each day, ILI Tracker is able to recognize the presence of a novel disease quickly. As we report in [34], the DUDE system recognized the outbreak of D68 on September 14, over two weeks later than ILI Tracker signaled it. Furthermore, by presenting a quantitative measure of how statistically unlikely the data for a particular day is—along with identifying specific patients—ILI Tracker provides a graded, actionable alert to clinicians.

## 8 Conclusions and Future Work

Results reported in this paper provide support that ILI Tracker was able to track well four modeled ILI-like diseases over a one-year period, relative to laboratory confirmed cases, and it was computationally efficient in doing so. The system was also able to detect a novel outbreak of EV-D68 early in an outbreak that occurred in Allegheny County in 2014, as well as clinically characterize that outbreak disease accurately. Detection was very efficient computationally. In general, ILI Tracker scales linearly in the number of diseases and in the number of findings per disease. Thus, it can be expanded to model many additional, known outbreak diseases and their findings.

In future work, we plan to extend ILI Tracker to model additional respiratory diseases, including adenovirus, enterovirus, and COVID-19. We also plan to expand the set of findings from less than the one hundred currently in use to many thousands that are encoded using UMLS concept unique identifiers (CUIs) [35]. In addition, we plan to evaluate ILI Tracker and its extensions on additional years of UPMC ED data from Allegheny County, Pennsylvania.

Our ultimate goal is to deploy an effective, free, open-source early-warning surveillance system for use in hospital emergency departments.

## Data Availability

Data used in this study includes patient-care records and is not generally available.

## 9 Acknowledgements

This work was supported by grant R01LM013509 (*Automated Surveillance of Overlapping Outbreaks and New Outbreak Diseases*) from the National Library of Medicine of the U.S. National Institutes of Health. Harry Hochheiser was supported by MIDAS grant U24GM132013. Ye Ye was partially supported by grant R00LM013383 from NIH/NLM. The content is solely the responsibility of the authors and does not necessarily represent the official views of the National Institutes of Health.

## 10 IRB Approval

The research protocol was approved by University of Pittsburgh IRB Study 20030193.

## Notes

### Competing Interest Statement

The authors have declared no competing interest.

### Author Declarations

Ethics committee/IRG of University of Pittsburgh gave ethical approval for this work.

## Bibliography

[1] Virginia Dato, Richard Shephard, and Michael M. Wagner. Outbreaks and investigations. In Handbook of Biosurveillance [36], pages 13–26.

[2] Michael M. Wagner, Louise S. Gresham, and Virginia Dato. Case detection, outbreak detection, and outbreak characterization. In Handbook of Biosurveillance [36], pages 27–50.

[3] Rita Velikina, Virginia Dato, and Michael M. Wagner. Governmental public health. In Handbook of Biosurveillance [36], pages 67–88.

[4] Michael M. Wagner, William R. Hogan, and Ron M. Aryel. The healthcare system. In Handbook of Biosurveillance [36], pages 89–110.

[5] Charles Brokopp, Eric Resultan, Harvey Holmes, and Michael M. Wagner. Laboratories. In Handbook of Biosurveillance [36], pages 129–142.

[6] C. Jessica E. Metcalf and Justin Lessler. Opportunities and challenges in modeling emerging infectious diseases. Science, 357:149–152, 2017.

[7] Edward C. Holmes, Andrew Rambaut, and Kristian G. Andersen. Pandemics: spend on surveillance, not prediction. Nature, 558:180–182, 2018.

[8] Fuchiang Tsui, Michael Wagner, Gregory Cooper, Jialan Que, Hendrik Harkema, John Dowling, Thomsun Sriburadej, Qi Li, Jeremy Espino, and Ronald Voorhees. Probabilistic case detection for disease surveillance using data in electronic medical records. Online Journal of Public Health Informatics, 3(3), 2011.

[9] Wendy W. Chapman and Henk Harkema. Identifying respiratory-related clinical conditions from ED reports with Topaz. Clinical Medicine and Research, 8(1), 2010.

[10] Gregory F. Cooper and Edward Herskovits. A Bayesian method for the induction of probabilistic networks from data. Machine Learning, 9:309–347, 1992.

[11] Marek J. Druzdzel. GeNIe: A development environment for graphical decision-analytic models. Proceedings of the 1999 Annual Symposium of the American Medical Informatics Association (AMIA-1999), 1999.

[12] J.A. Hanley and B.J. McNeil. The meaning and use of the area under a receiver operating characteristic (ROC) curve. Radiology, 143(1):29–36, 1982.

[13] Yongbo Liang, Derek Abbott, Newton Howard, Kenneth Lim, Rabab Ward, and Mohamed Elgendi. How effective is pulse arrival time for evaluating blood pressure? challenges and recommendations from a study using the mimic database. Journal of Clinical Medicine, 8(3):337, March 2019.

[14] Susan Prion and Katie Anne Haerling. Making sense of methods and measurement: Spearman-rho ranked-order correlation coefficient. Clinical Simulation in Nursing, 10(10):535–536, October 2014.

[15] Claire M. Midgley, Mary Anne Jackson, Rangaraj Selvarangan, George Turabelidze, Emily Obringer, Daniel Johnson, B. Louise Giles, Ajanta Patel, Fredrick Echols, M. Steven Oberste, W. Allan Nix, John T. Watson, and Susan I. Gerber. Severe respiratory illness associated with enterovirus D68 — Missouri and Illinois, 2014. Morbidity and Mortality Weekly Report (MMWR), 63(36), September 2014.

[16] Negar Aliabadi, Kevin Messacar, Daniel M. Pastula, Christine C. Robinson, Eyal Leshem, James Sejvar, W. Allan Nix, M. Steven Oberste, Daniel R. Feikin, and Samuel R. Dominguez. Enterovirus D68 infection in children with acute flaccid myelitis, Colorado, USA, 2014. Emerging Infectious Diseases, 22(8):1387–1394, 2016.

[17] Emilia Vynnycky and Richard G. White. An Introduction to Infectious Disease Modelling. Oxford University Press, 2010.

[18] M Biggerstaff, M Johansson, D Alper, LC Brooks, P Chakraborty, DC Farrow, S Hyun, S Kandula, C McGowan, and N Ramakrishnan. Results from the second year of a collaborative effort to forecast influenza seasons in the United States. Epidemics, 2018.

[19] JBS Ong, I Mark, C Chen, AR Cook, HC Lee, VJ Lee, RTP Lin, PA Tambyah, and LG Goh. Real-time epidemic monitoring and forecasting of H1N1-2009 using influenzalike illness from general practice and family doctor clinics in Singapore. PLoS One, 2010.

[20] J Shaman and A Karspeck. Forecasting seasonal outbreaks of influenza. Proceedings of the National Academy of Sciences, 109(50), 2012.

[21] LC Brooks, DC Farrow, S Hyun, RJ Tibshirani, and R Rosenfeld. Flexible modeling of epidemics with an empirical Bayes framework. PLoS Computational Biology, 11(8), 2015.

[22] EL Ray and NG Reich. Prediction of infectious disease epidemics via weighted density ensembles. PLoS Computational Biology, 14(2), 2018.

[23] DC Farrow, LC Brooks, S Hyun, RJ Tibshirani, DS Burke, and R Rosenfeld. A human judgment approach to epidemiological forecasting. PLoS computational biology, 13(3), 2017.

[24] Y Ye, F Tsui, M Wagner, JU Espino, and Q Li. Influenza detection from emergency department reports using natural language processing and Bayesian network classifiers. Journal of the American Medical Informatics Association, 21(5), 2014.

[25] GF Cooper, R Villamarin, F Tsui, N Millett, JU Espino, and MM Wagner. A method for detecting and characterizing outbreaks of infectious disease from clinical reports. Journal of Biomedical Informatics, 53, 2015.

[26] John M. Aronis, Nicholas E. Millett, Michael M. Wagner, Fuchiang Tsui, Ye Ye, Jeffrey P. Ferraro, Peter J. Haug, Per H. Gesteland, and Gregory F. Cooper. A Bayesian system to detect and characterize overlapping outbreaks. Journal of Biomedical Informatics, 73:171–181, 2017.

[27] Weng-Keen Wong and Andrew W. Moore. Classical time-series methods for biosurveillance. In Handbook of Biosurveillance [36], pages 217–234.

[28] Andrew W. Moore, Brigham Anderson, Kaustav Das, and Weng-Keen Wong. Combining multiple signals for biosurveillance. In Handbook of Biosurveillance [36], pages 235–242.

[29] Weng-Keen Wong, Andrew W. Moore, Gregory F. Cooper, and Michael M. Wagner. Bayesian network anomaly pattern detection for disease outbreaks. In Proceedings of the Twentieth International Conference on Machine Learning. AAAI Press, 2003.

[30] Howard Burkom, Yevgeniy Elbert, Christine Piatko, and Clay Fink. A term-based approach to asyndromic determination of significant case clusters. Online Journal of Public Health Informatics, 7(1), 2015.

[31] Mallory Nobles, Lana Deyneka, Amy Ising, and Daniel B. Neill. Identifying emerging novel outbreaks in textual emergency department data. Online Journal of Public Health Informatics, 7(1), 2015.

[32] Mallory Nobles, Ramona Lall, Robert Mathes, and Daniel Neill. Multidimensional semantic scan for pre-syndromic disease surveillance. Online Journal of Public Health Informatics, 11(1), 2019.

[33] Mallory Nobles, Ramona Lall, Robert W. Mathes, and Daniel B. Neill. Presyndromic surveillance for improved detection of emerging public health threats. Science Advances, 8(44), 2022.

[34] John M. Aronis, Jeffrey P. Ferraro, Per H. Gesteland, Fuchiang Tsui, Ye Ye, Michael M. Wagner, and Gregory F. Cooper. A Bayesian approach for detecting a disease that is not being modeled. PLoS ONE, February 2020.

[35] Olivier Bodenreider. The unified medical language system (umls): Integrating biomedical terminology. Nucleic Acids Research, 32:D267–D270, January 2004.

[36] Michael M. Wagner, Andrew W. Moore, and Ron M. Aryel. Handbook of Biosurveillance. Elsevier Academic Press, 2006.

